# Limited specificity of commercially available SARS-CoV-2 IgG ELISAs in serum samples of African origin

**DOI:** 10.1101/2020.09.15.20159749

**Authors:** Petra Emmerich, Carolin Murawski, Christa Ehmen, Ronald von Possel, Neele Pekarek, Lisa Oestereich, Sophie Duraffour, Meike Pahlmann, Nicole Struck, Daniel Eibach, Ralf Krumkamp, John Amuasi, Oumou Maiga-Ascofare, Raphael Rakotozandrindrainy, Danny Asogun, Yemisi Ighodalo, Simone Kann, Jürgen May, Egbert Tannich, Christina Deschermeier

## Abstract

Specific serological tests are mandatory for reliable SARS-CoV-2 seroprevalence studies but assay specificity may vary considerably between populations due to interference of immune responses to other pathogens. Here, we assess the false positive rates obtained with four commercially available IgG ELISAs in serum/plasma panels originating from three different African countries.

**Article summary line:** Several commercially available SARS-CoV-2 ELISAs show limited specificity when applied to serum panels of African origin.

---

By November 1^st^ 2020, 1,324,258 laboratory-confirmed SARS-CoV-2 infections and 29,785 deaths caused by COVID-19 have been reported from the WHO Africa region (1). Currently, community transmission is observed in almost all African countries including Tanzania, Ghana, Madagascar, Nigeria, Kenya, South Sudan, Burundi, and Uganda (1). To correctly determine the actual exposure of the population to SARS-CoV-2 and to draw reliable conclusions on morbidity and case fatality rates, highly sensitive and specific serological tests are mandatory.

Up to now, a plethora of ELISA tests for detection of anti-SARS-Cov-2 antibodies has been developed and commercialized (2, 3). Although performance data for several of these assays have been rapidly communicated by different laboratories (4-7), still only few reports are available on the applicability of these tests on African serum panels (8, 9). Here, assay specificity may be challenged by previous or current infections with other endemic pathogens like other coronaviruses (10), Dengue virus (11), or *Plasmodium spp. (12)*.

## THE STUDY

To assess the specificities of commercially available SARS-CoV-2 IgG ELISA tests in sample panels of different origin, *a priori* SARS-CoV-2 IgG negative sample panels (**Table 1**) collected from symptom-free donors before 2019 in Africa (Ghana, Madagascar, Nigeria), South America (Colombia) and Europe (Germany) were analyzed with the Euroimmun Anti-SARS-CoV-2-NCP IgG ELISA (Euroimmun, Germany), the Euroimmun Anti-SARS-CoV-2 IgG ELISA (Euroimmun, Germany), the EDI^™^ Novel Coronavirus COVID-19 IgG ELISA (Epitope Diagnostics, US), and the Mikrogen *recom*Well SARS-CoV-2 IgG ELISA (Mikrogen, Germany) (**Appendix Table 1**). Assays were performed and evaluated according to the manufacturers’ instructions.

**Table 1.**
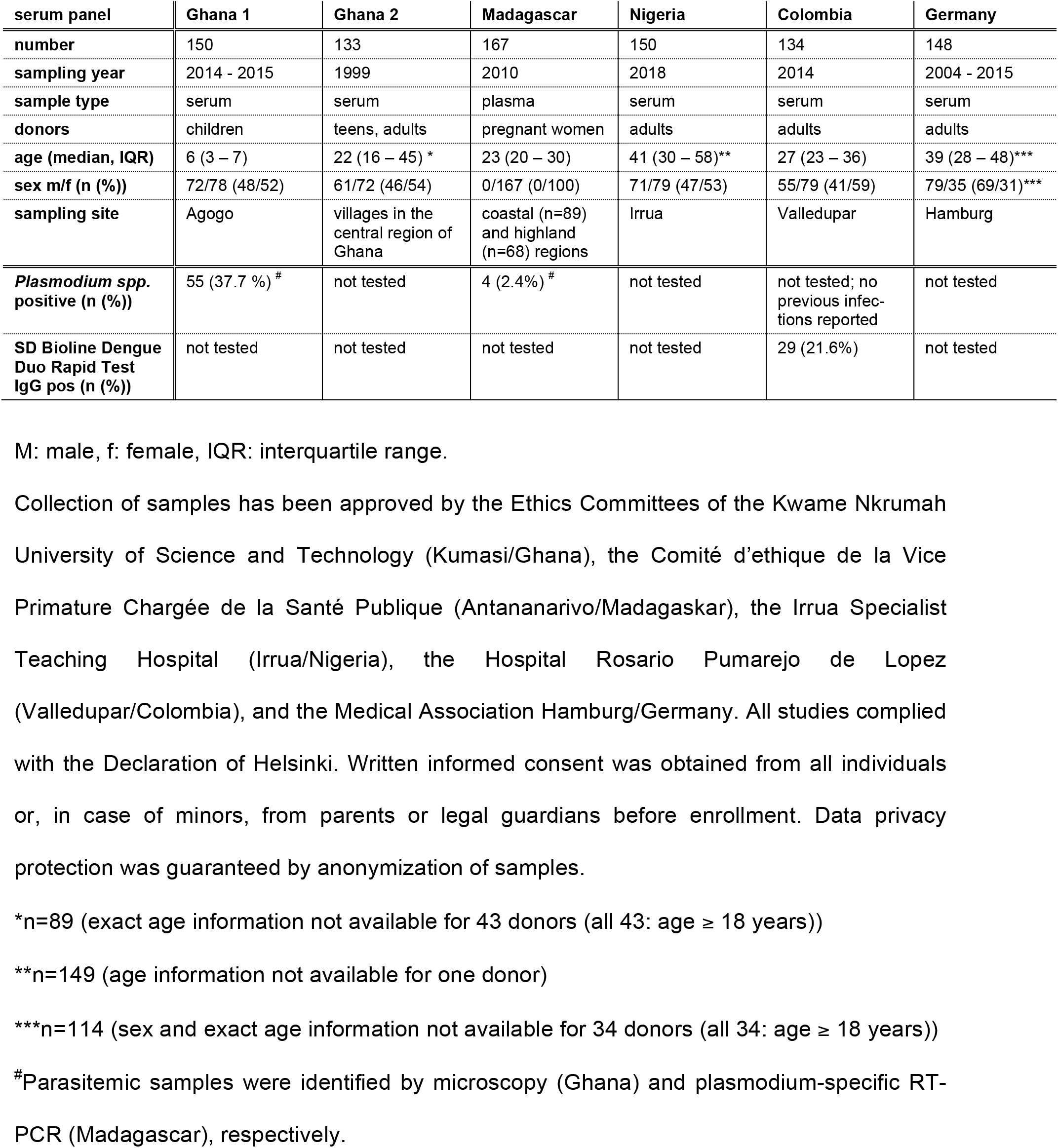
**Sample panels used in the study**

| serum panel | Ghana 1 | Ghana 2 | Madagascar | Nigeria | Colombia | Germany |
| --- | --- | --- | --- | --- | --- | --- |
| number | 150 | 133 | 167 | 150 | 134 | 148 |
| sampling year | 2014 - 2015 | 1999 | 2010 | 2018 | 2014 | 2004 - 2015 |
| sample type | serum | serum | plasma | serum | serum | serum |
| donors | children | teens, adults | pregnant women | adults | adults | adults |
| age (median, IQR) | 6 (3 – 7) | 22 (16 – 45) * | 23 (20 – 30) | 41 (30 – 58)** | 27 (23 – 36) | 39 (28 – 48)*** |
| sex m/f (n (%)) | 72/78 (48/52) | 61/72 (46/54) | 0/167 (0/100) | 71/79 (47/53) | 55/79 (41/59) | 79/35 (69/31)*** |
| sampling site | Agogo | villages in the central region of Ghana | coastal (n=89) and highland (n=68) regions | Irrua | Valledupar | Hamburg |
| *Plasmodium* spp. positive (n (%)) | 55 (37.7 %) # | not tested | 4 (2.4%) # | not tested | not tested; no previous infections reported | not tested |
| SD Bioline Dengue Duo Rapid Test IgG pos (n (%)) | not tested | not tested | not tested | not tested | 29 (21.6%) | not tested |

M: male, f: female, IQR: interquartile range.
Collection of samples has been approved by the Ethics Committees of the Kwame Nkrumah University of Science and Technology (Kumasi/Ghana), the Comité d'éthique de la Vice Primature Chargée de la Santé Publique (Antananarivo/Madagascar), the Irrua Specialist Teaching Hospital (Irrua/Nigeria), the Hospital Rosario Pumarejo de Lopez (Valledupar/Colombia), and the Medical Association Hamburg/Germany. All studies complied with the Declaration of Helsinki. Written informed consent was obtained from all individuals or, in case of minors, from parents or legal guardians before enrollment. Data privacy protection was guaranteed by anonymization of samples.
\*n=89 (exact age information not available for 43 donors (all 43: age ≥ 18 years))
\*\*n=149 (age information not available for one donor)
\*\*\*n=114 (sex and exact age information not available for 34 donors (all 34: age ≥ 18 years))

While IgG ELISA specificities where good to excellent for pre-COVID-19 sample panels originating from Colombia, Madagascar, and Germany, increased false positive rates were observed in *a priori* SARS-CoV-2 IgG negative sera from Ghana and Nigeria (**Figure 1, Table 2**). Thereby, the index values obtained with the Euroimmun NCP IgG ELISA and the EDI^™^ IgG ELISA, both employing recombinant nucleocapsid protein (NCP) as antigen, showed a clear correlation (**Appendix Figure 1A, 1C, 1E, 1G)**, while only 15 samples (Ghana: n=7, Nigeria: n=8), were concordantly classified as positive by both the NCP-based and the spike/S1-based Euroimmun IgG ELISA (**Appendix Figure 1B, 1D, 1F, 1H, Figure 2A)**. For all 15 sera, IgG immunofluorescence testing using SARS-CoV-2-infected Vero cells (13) was negative, and no or only weak inhibition of receptor binding was observed in a SARS-CoV-2 surrogate virus neutralizing test (sVNT, Genscript, US) (**Figure 2A**).

**Table 2.**
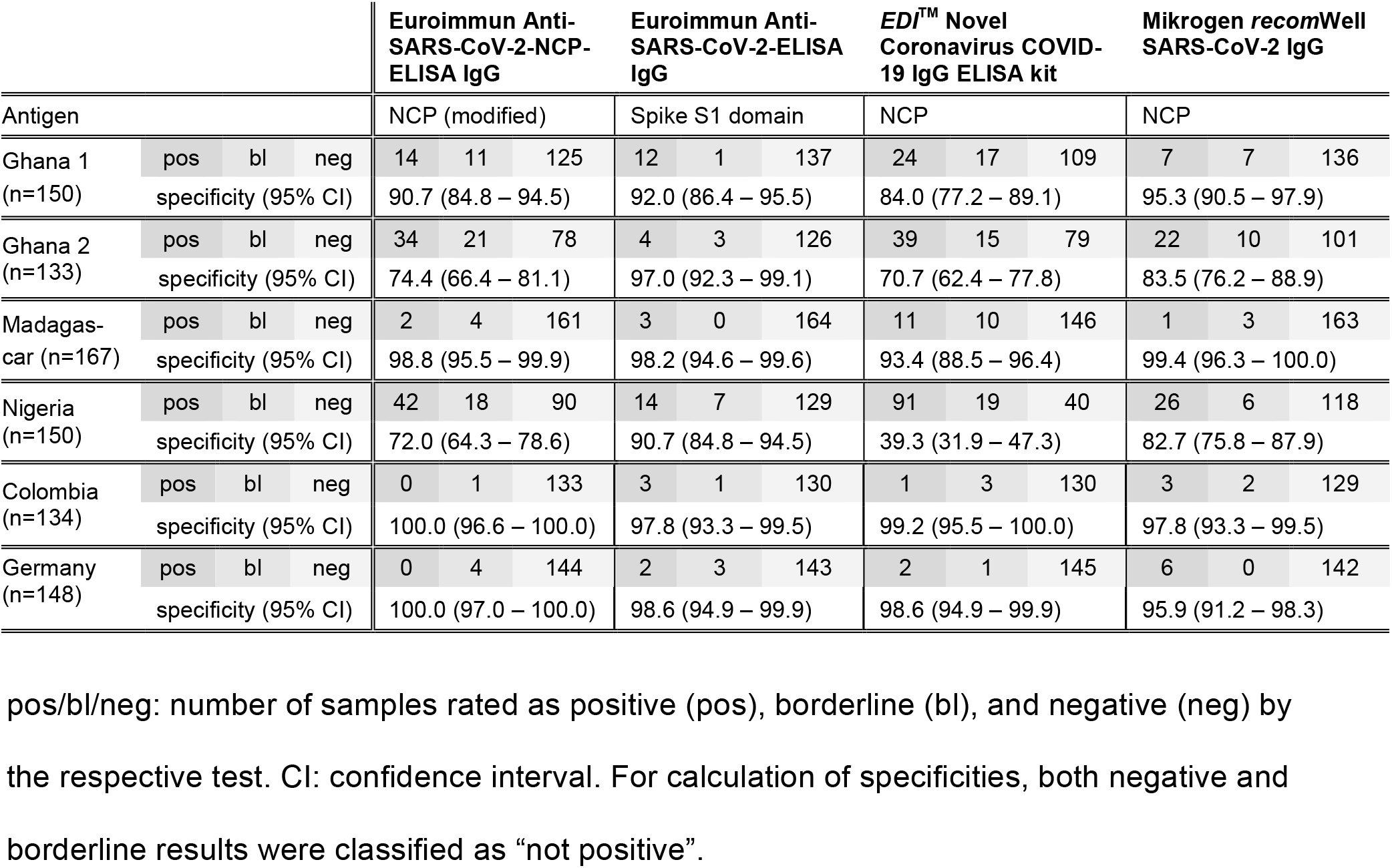
**SARS-CoV-2 IgG ELISA specificities**

**Figure 1.**
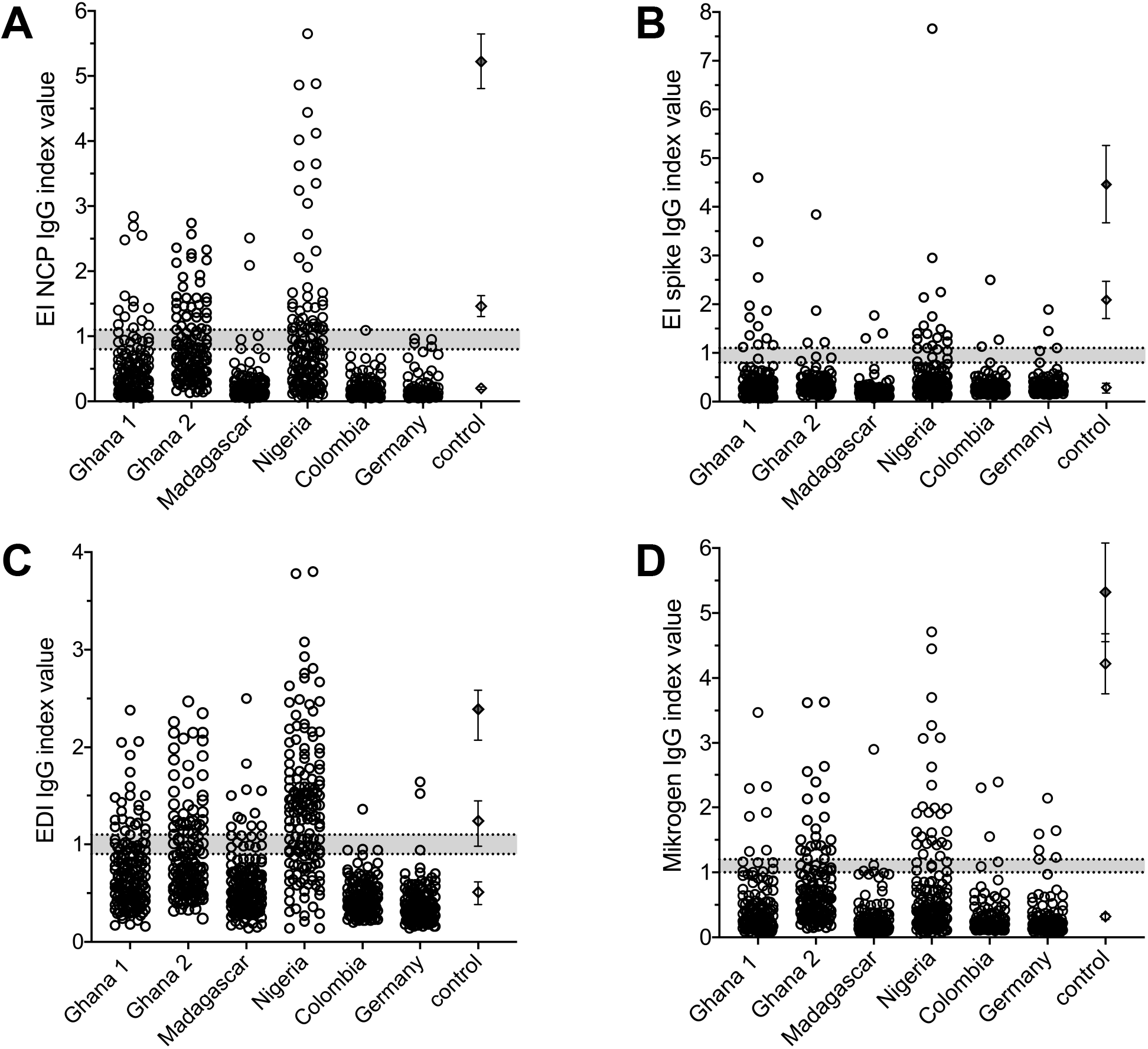
SARS-CoV-2 IgG ELISA results. Index values obtained for serum/plasma samples collected before 2019 in three different African countries (Ghana panel 1 (n=150), Ghana panel 2 (n=133), Madagascar (n=167), Nigeria (n=150)), Colombia (n=134), and Germany (n=148) with **(A)** the Euroimmun Anti-SARS-CoV-2-NCP IgG ELISA, **(B)** the Euroimmun Anti-SARS-CoV-2 IgG ELISA, **(C)** the EDI Novel Coronavirus COVID-19 IgG ELISA, and **(D)** the Mikrogen *recom*Well SARS-CoV-2 IgG ELISA. Dotted lines represent negative and positive cut-off values, respectively. Grey shading indicates index values rated as “borderline” according to the manufacturers’ instructions. Diamonds represent index values obtained for two IgG positive COVID-19 patient sera sampled on day 19 post onset of symptoms (dark grey: SARS-CoV-2 IgG IIFT titer 1:640, light grey: SARS-CoV-2 IgG IIFT titer 1:160) and one negative control serum; error bars represent standard deviation of n=13 (A, C) and n=14 (B, D) independent measurements, respectively.

**Figure 2.**
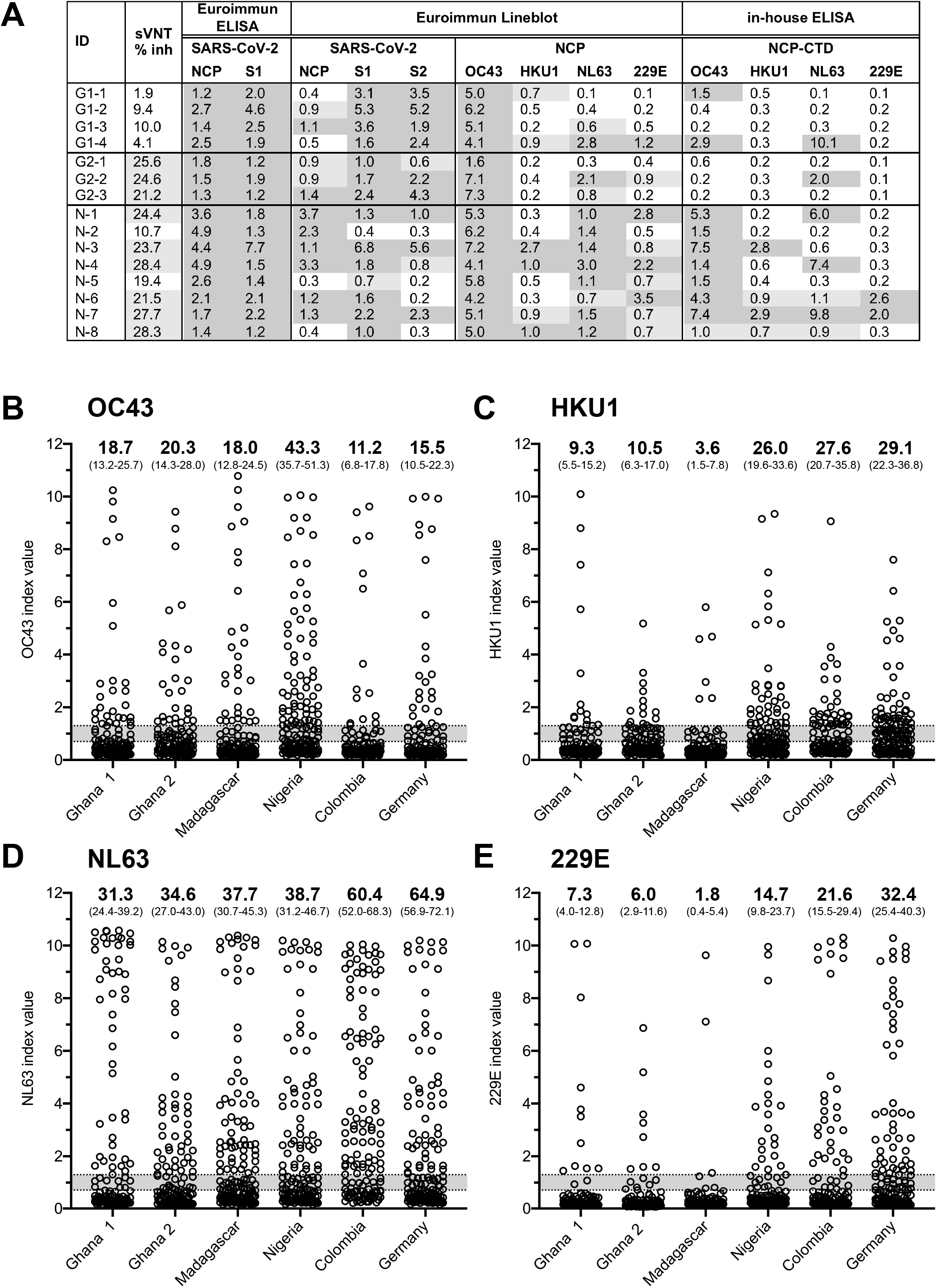
(A) ELISA, sVNT and line blot results for sera testing positive in both the Euroimmun spike/S1- and NCP-based IgG ELISA. Serum samples (Ghana 1 (G1): n=4, Ghana 2 (G2): n=3, Nigeria (N): n=8)) were tested using the SARS-CoV-2 sVNT (Genscript) and the Euroline Anti-SARS-CoV-2 Profile IgG (Euroimmun) according to the manufacturer’s instructions. Rating of index values (iv): Euroimmun ELISA: negative: iv < 0.8, borderline; 0.8 ≤ iv < 1.1, positive: iv ≥ 1.1; Line blot: negative: iv < 0.6, borderline: 0.6 ≤ iv < 1.0, positive: iv ≥ 1.0; in-house ELISA: negative: iv < 0.7, borderline: 0.7 ≤ iv < 1.3, positive: iv ≥ 1.3; sVNT: negative: % inhibition < 20.0, borderline; 20.0 ≤ % inh < 30.0, positive: % inh ≥ 30.0; CTD: C-terminal domain; dark grey fields: positive; light grey fields: borderline. **(B) - (E) Common cold CoVs ELISA results**. Index values obtained for the sample panels from Ghana (1: n=150, 2: n=133), Madagascar (n=167), Nigeria (n=150), Colombia (n=134), and Germany (n=148) using an in-house IgG ELISA protocol employing the C-terminal dimerization domain of **(A)** OC43 NCP, **(B)** HKU1 NCP, **(C)** NL63 NCP, and **(D)** 229E NCP as antigen. Bold numbers: % of samples for which an iv ≥ 1.3 was obtained, numbers in brackets: 95% confidence interval. Grey shading indicates ivs rated as “borderline” (0.7 ≤ iv < 1.3).

One possible cause of the observed limited specificity of SARS-CoV-2 IgG ELISAs may be cross-reaction with antibodies elicited by previous infections with other coronaviruses. To investigate this possibility, we assayed the complete sample panel for IgG antibodies binding to the C-terminal dimerization domains of the four common cold coronavirus NCPs using an in-house IgG ELISA protocol ((14), **Appendix Methods, Figure 2B-E**).

As can be expected due to the worldwide occurrence of the common cold coronaviruses, IgG antibodies interacting with the C-terminal domains of OC43, HKU1, NL63, and 229E NCP were detected in considerable fractions of all sample panels, including those for which good or excellent SARS-CoV-2 ELISA specificity has been observed (**Figure 2B-E)**. Nevertheless, the most challenging sample panel originating from Nigerian donors displayed the by far highest percentage of OC43-ELISA-reactive samples (43.3 %, **Figure 2B**). In a line blot (Euroline Anti-SARS-CoV-2 Profile IgG, Euroimmun), all 15 serum samples from Ghana (n=7) and Nigeria (n=8) showing up positive in both the spike/S1-based and the NCP-based Euroimmun IgG ELISA generated strong signals with OC43 NCP (**Figure 2A**). In addition, antibodies binding to SARS-CoV-2 spike S2 were found in 6/7 Ghanaian and 3/8 Nigerian samples (**Figure 2A**). 7/8 of the Nigerian sera, but only 2/7 of the Ghanaians samples tested positive in the in-house OC43 ELISA employing a truncated NCP lacking the N-terminal domain as antigen. Therefore, at least some of the observed false positive signals could be due to previous infections with other coronaviruses sharing B cell epitopes with both OC43 and SARS-CoV-2.

Recently, Lustig *et al. (11)* reported false positive results of the Euroimmun Anti-SARS-CoV-2 IgA and IgG ELISAs in sera originating from donors with acute or past dengue virus (DENV) infection. In our study, 29 (21.6%) of the 134 Colombian donors tested positive in the SD Bioline Dengue Duo IgG Rapid Test (Alere/Abbott, USA), indicating a high titer of anti-DENV IgG antibodies (15). Indeed, two of the three Colombian sera testing positive in the spike-based Euroimmun Anti-SARS-CoV-2 ELISA (but negative in the nucleoprotein-based SARS-CoV-2 ELISAs) belonged to this subgroup.

In addition, hypergammaglobulinemia resulting from polyclonal B-cell activation induced by pathogens like plasmodium can challenge assay specificity as has already been shown for a commercially available ZIKV IgG ELISA (12). Information about *Plasmodium* parasitemia was only available for one of the Ghanaian (55/150 samples with microscopically detectable parasitemia) and the Madagascan (4/167 Plasmodium-PCR positive samples) panels. Here, a reduced specificity in Ghanaian parasitemic vs. non-parasitemic samples was observed for the Euroimmun Anti-SARS-CoV-2-NCP-IgG ELISA (83.6% (95% CI: 71.5%-91.4%) vs. 94.7% (95% CI: 88.0%-98.0%), p = 0.0387), but not for the other SARS-CoV-2 IgG ELISAs (**Appendix Table 2**).

## LIMITATIONS OF THE STUDY

Our study has some major limitations that have to be acknowledged when interpreting the presented data. First of all, sample panels originating from previous studies with different scientific objectives had to be used for the analysis; therefore, the panels do not reflect representative cross-sections of the respective countries’ populations and also comparability between sample panels is limited due to differences in age and sex of donors. Although in total 600 African samples have been analyzed, the number of samples per country giving rise to false positive SARS-CoV-2 IgG assay results is still relatively small, impeding statistical analyses. Furthermore, limited accessible sample volumes prevented us from performing material-intensive assays as the SD Bioline Dengue Duo IgG Rapid Test or *Plasmodium*-specific RT-PCR for the complete panel.

## CONCLUSIONS

In accordance with evaluation studies recently published by other authors (4-6), the commercially available SARS-CoV-2 IgG ELISAs displayed a good to excellent specificity when applied to serum panels originating from European donors. In contrast, significantly increased false positive rates were observed in African pre-COVID-19 serum panels originating from Ghana and Nigeria. A similar result has been found recently for serum samples from febrile patients from Benin (9). Possible causes for this observation are cross-reactive antibodies elicited by previous infections with endemic viruses (in particular other coronaviruses), polyclonal B-cell activation induced e.g. by plasmodium infection, or a combination thereof.

Based on these findings, the following recommendations should be considered when planning and performing SARS-CoV-2 seroprevalence studies (not only) in Africa: 1) Prior to performance of seroprevalence studies, carefully assess background/false positive signals obtained with the chosen serological test(s) in the target population (using *a priori* SARS-CoV-2 IgG negative serum samples which were stocked before 2019). Be aware of potentially interfering/cross-reacting endemic pathogens and carefully interpret SARS-CoV-2 IgG test results in this context. 2) If necessary, combine information from two independent serological tests employing different antigens. 3) Re-evaluate samples generating a positive ELISA result by SARS-CoV-2 neutralization testing.

Further studies will be necessary to assess sensitivity of the commercially available assays in detecting anti-SARS-CoV2 IgG antibodies induced by SARS-CoV-2 infection in African COVID-19 patients and to optimize assay specificity in sera from African donors.

## Supporting information

STARD Flowchart

STARD Checklist

## Data Availability

All data relevant for the study are presented in the manuscript and its appendix.

## ACKNOWLEDGMENTS

The authors thank M. Panning for providing common cold coronavirus RNAs. J. Hansen is thanked for expert technical assistance.

## AUTHOR BIOGRAPHY

Dr. Emmerich is a laboratory group leader at the Bernhard Nocht Institute for Tropical Medicine in Hamburg, Germany. Her research is focused on diagnostics of viral infectious diseases.

## REFERENCES

1. WHO. COVID-19 Weekly Epidemiological Update, 01Nov2020

2. FIND. https://www.finddx.org/covid-19/sarscov2-eval-immuno/

3. University JH. https://www.centerforhealthsecurity.org/resources/COVID-19/serology/Serology-based-tests-for-COVID-19.html. 2020.

4. Geurts van Kessel CH, Okba NMA, Igloi Z, Bogers S, Embregts CWE, Laksono BM, et al. An evaluation of COVID-19 serological assays informs future diagnostics and exposure assessment. Nature communications. 2020;11(1):3436.

5. Kruttgen A, Cornelissen CG, Dreher M, Hornef M, Imohl M, Kleines M. Comparison of four new commercial serologic assays for determination of SARS-CoV-2 IgG. Journal of clinical virology : the official publication of the Pan American Society for Clinical Virology. 2020;128:104394.

6. Pflüger LS, Bannasch JH, Brehm TT, Pfefferle S, Hoffmann A, Nörz D., et al. Clinical evaluation of five different automated SARS-CoV-2 serology assays in a cohort of hospitalized COVID-19 patients. Journal of Clinical Virology. 2020.

7. Weidner L, Gansdorfer S, Unterweger S, Weseslindtner L, Drexler C, Farcet M, et al. Quantification of SARS-CoV-2 antibodies with eight commercially available immunoassays. Journal of clinical virology : the official publication of the Pan American Society for Clinical Virology. 2020;129:104540.

8. Chibwana MG, Jere KC, Kamn’gona R, Mandolo J, Katunga-Phiri V, Tembo D, et al. High SARS-CoV-2 seroprevalence in health care workers but relatively low numbers of deaths in urban Malawi. MedRxiv preprint, https://doi.org/10.1101/2020.07.30.20164970. 2020.

9. Yadouleton A, Sander A, Moreira-Soto A, Tchibozo C, Hounkanrin G, Badou Y, et al. Limited specificity of serologic tests for SARS-CoV-2 antibody detection, Benin, Western Africa. medRxiv preprint, https://doi.org/10.1101/2020.06.29.20140749. 2020.

10. Tso FY, Lidenge SJ, Pena PB, Clegg AA, Ngowi JR, Mwaiselage J., et al. High prevalence of pre-existing serological cross-reactivity against SARS-CoV-2 in sub-Saharan Africa. International Journal of Infectious Diseases. https://doi.org/10.1016/j.ijid.2020.10.104. 2020

11. Lustig Y, Keler S, Kolodny R, Ben-Tal N, Atias-Varon D, Shlush E, et al. Potential antigenic cross-reactivity between SARS-CoV-2 and Dengue viruses. Clin Infect Dis. 2020.

12. Van Esbroeck M, Meersman K, Michiels J, Arien KK, Van den Bossche D. Letter to the editor: Specificity of Zika virus ELISA: interference with malaria. Euro surveillance : bulletin Europeen sur les maladies transmissibles = European communicable disease bulletin. 2016;21(21).

13. Reisinger EC, von Possel R, Warnke P, Geerdes-Fenge HF, Hemmer CJ, Pfefferle S, et al. Screening of Mothers in a COVID-19 Low-Prevalence Region: Determination of SARS-CoV-2 Antibodies in 401 Mothers from Rostock by ELISA and Confirmation by Immunofluorescence. Deutsche medizinische Wochenschrift. 2020.

14. Schmitz H, Gabriel M, Emmerich P. Specific detection of antibodies to different flaviviruses using a new immune complex ELISA. Med Microbiol Immunol. 2011;200(4):233–9.

15. Blessmann J, Winkelmann Y, Keoviengkhone L, Sopraseuth V, Kann S, Hansen J, et al. Assessment of diagnostic and analytic performance of the SD Bioline Dengue Duo test for dengue virus (DENV) infections in an endemic area (Savannakhet province, Lao People’s Democratic Republic). PLoS One. 2020;15(3):e0230337.

