## Appendix for "Limited specificity of commercially available SARS-CoV-2 IgG ELISAs in serum samples of African origin"

#### Appendix Methods

IgG antibodies directed against common cold coronaviruses OC43, HKU1, NL63, and 229E were detected using an immune complex binding ELISA platform technology patented by BNITM (patent number EP2492689). Briefly, diluted patient serum/plasma samples were co-incubated together with a biotinylated recombinant antigen for 24h at 4°C in a microwell plate coated with a recombinant IgG immune complex specific capture molecule. Following a washing step, the bound IgG/antigen immune complexes were visualized by subsequent application of horseradish peroxidase (HRP)-labeled streptavidin and the colorimetric HRP substrate tetramethylbenzidine (TMB). After stopping the enzymatic reaction, the assay result was generated by measuring the optical density of the solution in the well at 450/620 nm.

As antigens, bacterially expressed, N-terminally truncated common cold CoV nucleoproteins comprising the protein's dimerization domain were used (OC43 (AIL49389.1): NPCΔ258, HKU1 (AGT17773.1): NPCΔ256/S290F, NL63 (AFO70495.1): NPCΔ222, 229E (NP_073556.1): NPCΔ235). Similar constructs have been shown to enable sensitive and specific detection of IgG antibodies directed against SARS-CoV-1 (Yu *et al.*, 2005) and OC43 (Blanchard *et al.*, 2011).

An assay cut-off of OD450-OD620 = 0.3 was determined by comparison of the ELISA results with the results obtained with a commercially available lineblot (Euroline Anti-SARS-CoV-2 Profile IgG, Euroimmun) for 32 German healthy blood donors. Index values (iv) were calculated by dividing the measured OD450-OD620 values by 0.3; samples were classified as negative ($iv < 0.7$), borderline ($0.7 \leq iv < 1.3$), or positive ($iv \geq 1.3$).

#### References to Appendix Methods:


**Yu F**, Le MQ, Inoue S, Thai HT, Hasebe F, Parquet MDC, and Morita K. Evaluation of inapparent nosocomial severe acute respiratory syndrome coronavirus infection in Vietnam by use of highly specific recombinant truncated nucleocapsid protein-based enzyme-linked immunosorbent assay. *Clin. Diagn. Lab. Immunol.* 2005; 12:848–854.

**Blanchard EG**, Miao C, Haupt TE, Anderson LJ, Haynes LM. Development of a recombinant truncated nucleocapsid protein based immunoassay for detection of antibodies against human coronavirus OC43. *J. Virol. Methods* 2011; 177(1):100-6.

#### Appendix Table 1. Commercially available ELISA kits used in the study

| | Euroimmun Anti-SARS-CoV-2-NCP-ELISA IgG | Euroimmun Anti-SARS-CoV-2-ELISA IgG | *EDI*™ Novel Coronavirus COVID-19 IgG ELISA kit | Mikrogen *recom*Well SARS-CoV-2 IgG |
|---|---|---|---|---|
| **manufacturer** | Euroimmun AG<br>Lübeck, Germany | Euroimmun AG<br>Lübeck, Germany | Epitope Diagnostics, Inc.<br>San Diego, US | Mikrogen GmbH<br>Neuried, Germany |
| **status** | CE-IVD | CE-IVD, FDA EUA | CE-IVD | CE-IVD |
| **antibody isotype** | IgG | IgG | IgG | IgG |
| **test format** | 96 well microplate | 96 well microplate | 96 well microplate | 96 well microplate |
| **test principle** | indirect ELISA | indirect ELISA | indirect ELISA | indirect ELISA |
| **antigen** | NCP (modified) | spike (S1 domain) | NCP | NCP |
| **sample dilution** | 1:101 | 1:101 | 1:101 | 1:101 |
| **interpretation index value (iv)** | negative: iv < 0.8<br>borderline; 0.8 ≤ iv < 1.1<br>positive: iv ≥ 1.1 | negative: iv < 0.8<br>borderline; 0.8 ≤ iv < 1.1<br>positive: iv ≥ 1.1 | negative: iv ≤ 0.9<br>borderline; 0.9 < iv < 1.1<br>positive: iv ≥ 1.1 | negative: iv < 1.0<br>borderline; 1.0 ≤ iv < 1.2<br>positive: iv ≥ 1.2 |
| **references** | (*7*) | (*4-7*) | (*5*) | (*5*) |

CE: Conformité Européenne, IVD: *in vitro* diagnostics; FDA: Food and Drug Administration; EUA: Emergency Use Authorization; NCP: nucleocapsid protein; IFU: Instructions for Use

#### Appendix Table 2. SARS-CoV-2 IgG ELISA specificities for donor subgroups from Ghana with and without *Plasmodium* parasitemia

| | | | | Euroimmun Anti-SARS-CoV-2-NCP-ELISA IgG | | | Euroimmun Anti-SARS-CoV-2-ELISA IgG | | | *EDI*™ Novel Coronavirus COVID-19 IgG ELISA kit | | | Mikrogen *recom*Well SARS-CoV-2 IgsG | | |
|---|---|---|---|---|---|---|---|---|---|---|---|---|---|---|---|
| parasitemic (n = 55) | pos | bl | neg | 9 | 6 | 40 | 6 | 1 | 48 | 12 | 9 | 34 | 1 | 4 | 50 |
| | specificity (95% CI) | | | 83.6 (71.5 – 91.4) | | | 89.1 (77.8 – 95.3) | | | 78.2 (65.5 – 87.2) | | | 98.2 (89.5 – 100.0) | | |
| not parasitemic (n = 95) | pos | bl | neg | 5 | 5 | 85 | 6 | 0 | 89 | 12 | 8 | 75 | 6 | 3 | 86 |
| | specificity (95% CI) | | | 94.7 (88.0 – 98.0) | | | 93.7 (86.6 – 97.3) | | | 87.4 (79.1 – 92.8) | | | 93.7 (86.6 – 97.3) | | |
| | | | p value | 0.0387 | | | 0.3578 | | | 0.1675 | | | 0.4235 | | |

pos/bl/neg: number of samples rated as positive (pos), borderline (bl), and negative (neg) by the respective test. CI: confidence interval. For calculation of specificities, both negative and borderline results were classified as “not positive”. P values were calculated using Fisher’s exact test (2 x 2 contingency table, pos versus “not positive”).

#### LEGEND

**Appendix Figure 1. Correlation of SARS-CoV-2 IgG ELISA results.** Index values obtained for the Ghanaian (**A, B, C, D**), Madagascan (**E, F**), and Nigerian (**G, H**) samples with the Euroimmun Anti-SARS-CoV-2-NCP IgG ELISA and the nucleoprotein-based EDI Novel Coronavirus COVID-19 IgG ELISA **(A, C, E, G)** or the spike-based Euroimmun Anti-SARS-CoV-2 IgG ELISA **(B, D, F, H)**. Dotted lines represent negative and positive cut-off values, respectively. Grey shading indicates index values rated as "borderline" according to the manufacturers' instructions. Numbers represent test specificity obtained upon combination of the two respective assays (only samples testing positive in both assays are classified as positive) and 95% CI.

### Appendix Figure 1

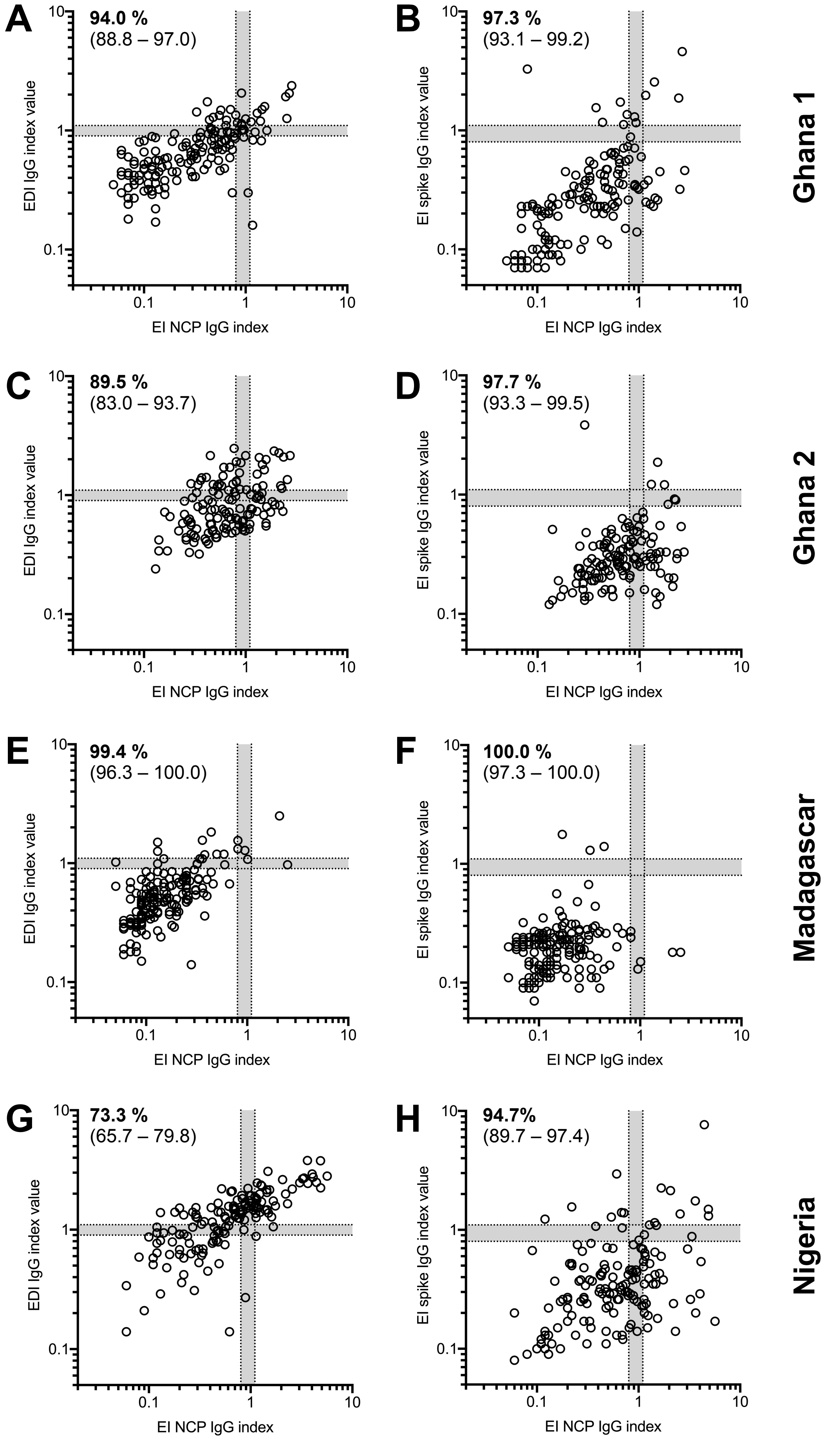
