## Supplementary figures and images for "Limited specificity of commercially available SARS-CoV-2 IgG ELISAs in serum samples of African origin"

### STARD Flowchart

#### STARD Flow Chart

**See also Appendix Table 1**

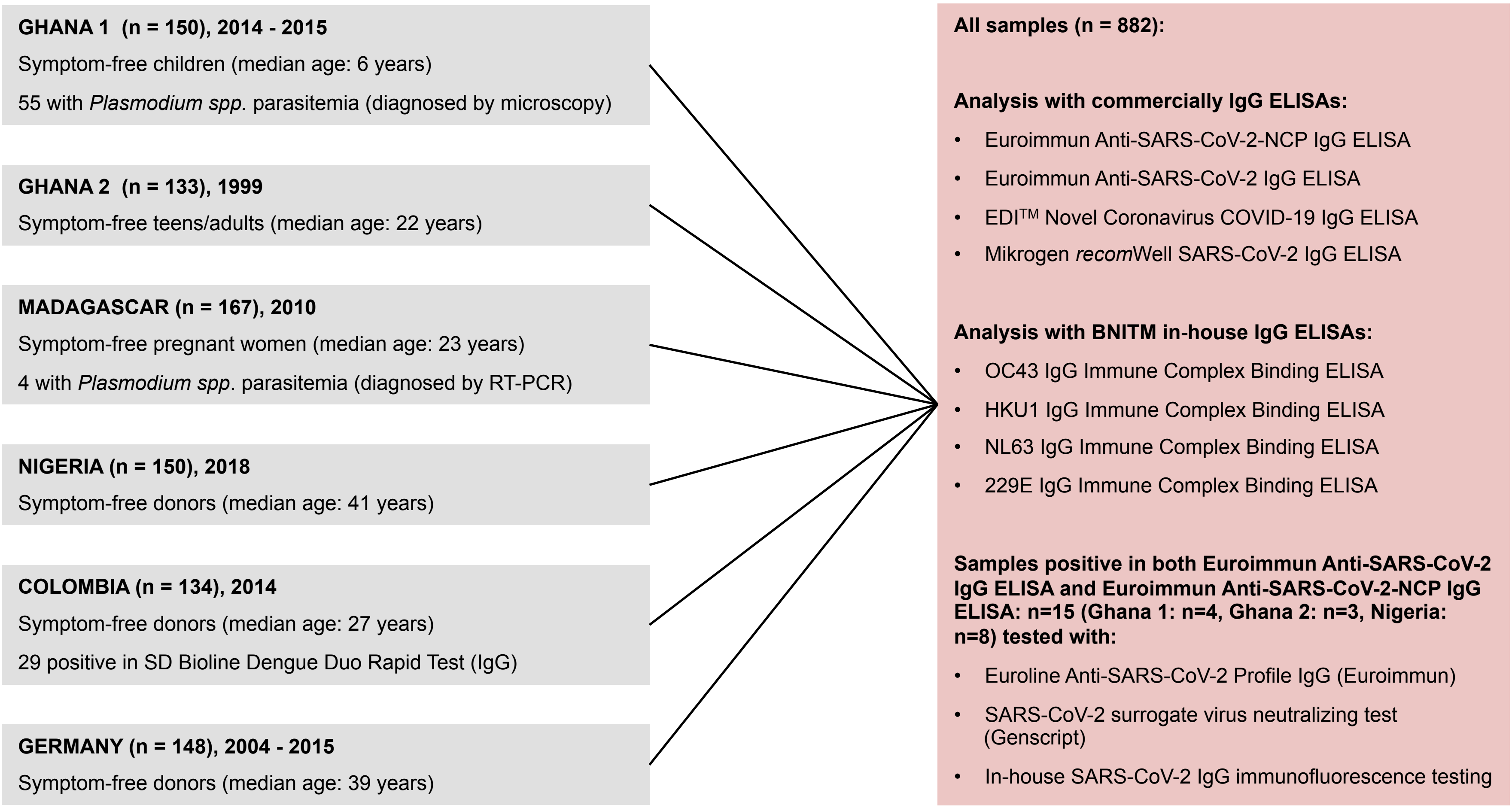
